# Brain Atrophy in Spinocerebellar Ataxia Type 1 (SCA1) across the Disease Course: MRI Volumetrics from ENIGMA-Ataxia

**DOI:** 10.64898/2026.04.22.26351550

**Authors:** Jason W. Robertson, Isaac Adanyeguh, Tetsuo Ashizawa, Benjamin Bender, Fernando Cendes, Giulia Coarelli, Andreas Deistung, Stefano Diciotti, Alexandra Durr, Jennifer Faber, Marcondes C. França, Sophia L Göricke, Marina Grisoli, James M. Joers, Thomas Klockgether, Christophe Lenglet, Caterina Mariotti, Alberto R. M. Martinez, Chiara Marzi, Mario Mascalchi, Anna Nigri, Gülin Öz, Henry Paulson, Maria J. Rakowicz, Kathrin Reetz, Thiago JR Rezende, Lidia Sarro, Ludger Schols, Matthis Synofzik, Dagmar Timmann, Sophia I Thomopoulos, Paul M Thompson, Bart van de Warrenburg, Carlos R. Hernandez-Castillo, Ian H. Harding

**Affiliations:** Faculty of Computer Science, Dalhousie University, Halifax, Canada Dalhousie University; Center for Magnetic Resonance Research and Department of Radiology, University of Minnesota Medical School, Minneapolis, Minnesota, USA; Weill Cornell Medicine at Houston Methodist Research Institute, Houston, TX 77030, USA; Department of Diagnostic and Interventional Neuroradiology, University Hospital Tübingen, Tübingen, Germany; Department of Neurology, School of Medical Sciences, University of Campinas (UNICAMP), Campinas, SP, Brazil; Brazilian Institute of Neuroscience and Neurotechnology (BRAINN), School of Medical Sciences, University of Campinas (UNICAMP), Campinas, SP, Brazil; Sorbonne Université, Paris Brain Institute (ICM), Pitié-Salpêtrière Hospital, AP-HP, INSERM, CNRS, University Hospital Pitié-Salpêtrière, Paris, France; University Clinic and Outpatient Clinic for Radiology, Department for Radiation Medicine, University Hospital Halle (Saale), University Medicine Halle, Halle (Saale), Germany; Department of Electrical, Electronic, and Information Engineering “Guglielmo Marconi”, University of Bologna, Cesena, Italy; German Center for Neurodegenerative Diseases (DZNE), Bonn, Germany; Department of Parkinson’s Disease, Sleep and Movement Disorders, Center of Neurology, University Hospital Bonn, Bonn, Germany; Department of Neuroradiology, University Hospital Bonn, Bonn, Germany; Institute of Diagnostic and Interventional Radiology and Neuroradiology and Center for Translational Neuro- and Behavioral Sciences (C-TNBS), Essen University Hospital, University of Duisburg-Essen, Essen, Germany; Department of Neuroradiology, Fondazione IRCCS Istituto Neurologico Carlo Besta, Milan, Italy; Department of Statistics, Computer Science, Applications “Giuseppe Parenti”, University of Florence, Florence; Department of Clinical and Experimental Biomedical Sciences “Mario Serio”, University of Florence, Florence, Italy; Department of Neurology, University of Michigan, Ann Arbor, Michigan, USA; Department of Clinical Neurophysiology, Institute of Psychiatry and Neurology, Warsaw, Poland; Department of Neurology, RWTH Aachen University, Aachen, Germany; JARA-BRAIN Institute Molecular Neuroscience and Neuroimaging, Research Center Jülich GmbH, Jülich, Germany; Department of Neurosciences, School of Medicine, University of São Paulo at Ribeirão Preto – USP-RP; Department of Neurodegenerative Diseases, Hertie Institute for Clinical Brain Research, University of Tübingen, Germany; German Center for Neurodegenerative Diseases (DZNE), Tübingen, Germany; Department of Neurology and Center for Translational Neuro- and Behavioral Sciences (C-TNBS), Essen University Hospital, University of Duisburg-Essen, Essen, Germany; Imaging Genetics Center, Mark and Mary Stevens Neuroimaging and Informatics Institute, Keck School of Medicine, University of Southern California, Marina del Rey, CA, USA; Department of Neurology, Donders Institute for Brain, Cognition, and Behaviour, Radboud University Medical Center, Nijmegen, The Netherlands; Queensland Institute of Medical Research (QIMR Berghofer), Brisbane, QLD, Australia; School of Translational Medicine, Monash University, Melbourne, VIC, Australia

## Abstract

**Objective:** Spinocerebellar ataxia type 1 (SCA1) is a rare, inherited neurodegenerative disease characterised by progressive deterioration of motor and cognitive function. Here, we illustrate the pattern and evolution of brain atrophy in people with SCA1 using a large multisite dataset.

**Methods:** Structural magnetic resonance imaging data from SCA1 (*n*=152) and healthy control (*n*=131) participants from seven sites and two consortia were analyzed using voxel-based morphometry. Cross-sectional stratification and correlations were undertaken with ataxia severity and duration to profile disease evolution. Cerebrocerebellar structural covariance analysis was used to understand the relationship between cerebral and cerebellar tissue atrophy.

**Results:** Atrophy in SCA1 first manifests in the lower brainstem and cerebellar white matter (WM), before progressing to the pons, anterior cerebellum, and cerebellar lobule IX. The midbrain and peri-thalamic WM and the remainder of the cerebellar cortex are then affected, with preferential involvement of specific motor and cognitive areas. Finally, degeneration in the striatum and cerebral WM corresponding to the corticospinal tract become apparent. Atrophy and correlations with ataxia severity are most pronounced in the cerebellar WM and pons. Structural covariance analysis showed reduced correlations between cerebellar and cerebral WM volume in SCA1 participants.

**Interpretation:** Cross-sectional stratification of a large SCA1 cohort by ataxia severity indicates a pattern of atrophy spread across the brainstem, cerebellum, and subcortical grey and white matter. Ongoing volume loss throughout the disease course is most evident in a core set of infra-tentorial brain regions. Atrophy of cerebellum spans both motor and cognitive functional zones. Cerebellar degeneration is not directly mirrored by downstream effects in the cerebrum.

## Introduction

Spinocerebellar ataxia type 1 (SCA1) is an autosomal dominant inherited, progressive neurodegenerative disorder.^1,2^ Gait ataxia is typically the first symptom to manifest, most commonly in the third or fourth decade of life, alongside or followed by limb ataxia, dysarthria and dysphagia, nystagmus, peripheral neuropathy, pyramidal tract signs, and cerebellar cognitive affective syndrome (CCAS). SCA1 is caused by a CAG triplet expansion in the *ATXN1* gene on the short arm of chromosome 6^1^, resulting in a mutated ataxin-1 protein having a toxic gain of function.^2^ This, in turn, produces a neurodegenerative effect that includes Purkinje cell death; atrophy of the pons, inferior olive, and deep cerebellar nuclei; and degeneration in the spinal cord, peripheral nerves, and cerebello-thalamo-cortical and basal ganglia-thalamo-cortical loops.^1,2^

Non-invasive imaging techniques such as magnetic resonance imaging (MRI) have been used to describe the brain changes associated with SCA1. Voxel-based morphometry (VBM) and cerebellar parcellation analyses have consistently reported widespread atrophy in the cerebellar grey matter (GM) and white matter (WM).^3–11^ Volume loss has also been observed across the entirety of the brainstem, with strongest effects in the pons, as well as in the thalamus, basal ganglia, and cerebellar peduncles.^3–6,8,9,11– 13^ The most robust volume differences in the brainstem and cerebellar WM are detectable even prior to the onset of ataxia symptoms.^8,9^ Thinning of the cerebral cortex, most prominently in the frontal lobes, has also been reported.^6,8^ Diffusion MRI, which focuses specifically on WM, has found decreases in microstructural integrity in the cerebellar and cerebral peduncles, medial and lateral lemnisci, spinothalamic and corticospinal tracts, corona radiata, and pontine crossing tracts.^3,6,9,12,14^ Many of these findings correlate in turn with disease duration and/or ataxia severity.^3,4,8,9,11,12,14^

SCA1 studies to date have primarily relied on cross-sectional and longitudinal assessments over intervals of up to 1 year in modestly-sized cohorts. These studies have been formative in identifying potential biomarkers of disease expression and progression, particularly in highlighting the value of pons volume, diffusion measures, and spectroscopy as sensitive longitudinal markers of disease progression.^8,9,11,12,15^ Despite this, understanding of how the magnitude and profile of atrophy evolve across the whole brain over the disease course remains limited. The relative rarity of SCA1 has been a particular barrier to collecting large MRI research cohorts that allow for robust inferences.

The principal objective of this study was to characterise SCA1 atrophy patterns and evolution using a multi-site dataset aggregated by the ENIGMA-Ataxia working group, including pre-ataxic individuals. In addition to describing the overall pattern of volume loss in individuals with SCA1 relative to controls, we undertake cross-sectional analyses of subgroups defined by different levels of ataxia severity to characterize the whole-brain spatial evolution of this disease. We additionally profile how SCA1 atrophy maps across different functional zones within the cerebellum.^16^ Finally, we evaluate the relationship between cerebral and cerebellar atrophy using cerebrocerebellar covariance analysis.

## Methods

### Participants and data

This study used a cross-sectional design with retrospectively collected data aggregated through the ENIGMA-Ataxia working group. Structural images, demographics, and clinical data were collected from SCA1 participants (*n* = 152) and healthy controls (CONT; *n* = 131) from seven sites (Campinas, Brazil; Essen/Halle, Germany; Florence, Italy; Milan, Italy; Minnesota, USA; Paris, France; and Tübingen, Germany) and two collaborating consortia (EUROSCA and READISCA) in accordance with their respective ethics and governance bodies. All data were anonymized and given new subject identifiers prior to aggregation. All procedures were approved by both the Monash University Human Research Ethics Committee and the Dalhousie University Research Ethics Board.

Participants with SCA1 were included based on: (1) at least 39 CAG repeats in the long allele of the *ATXN1* gene^1^, including individuals without clinically manifest ataxic symptoms, and/or (2) a genetically confirmed family history of SCA1 and clinical manifestation of disease-consistent ataxia. Age at ataxia onset was recorded based on self-report of the participant’s first ataxic symptoms. Ataxia severity at the time of MRI collection was determined using the Scale for Assessment and Rating of Ataxia (SARA; 6 sites)^17^, the International Cooperative Ataxia Rating Scale (ICARS; 1 site)^18^, or both (1 site). For analytical consistency, ICARS scores were converted to SARA based on the linear regression reported by Rummey *et al*.^*19*^ To corroborate the validity of this approach, converted scores were also calculated for 15 participants from the site that collected both metrics; the test-retest reliability between the estimated and actual SARA scores was very high (ICC=0.96).

A whole-brain, high-resolution T_1_-weighted structural MRI image was collected for each subject. Imaging protocols varied by site (**Supplementary Table S1**), but were consistent across patients and controls at each site. One site (Milan) collected data for both cohorts using two different protocols; these were treated as separate sites to best control for site effects.

Quality control of the imaging data (see “Image Processing”, below) resulted in exclusion of 12 SCA1 and 3 CONT, providing an initial dataset of 140 SCA1 and 128 CONT participants. Subsequently, sites without site-specific control data (EUROSCA: n=17 SCA1) and sites with small samples (READISCA Skyra subcohort: 4 SCA1, 2 CONT) were excluded due to an inability to control adequately for site-specific confounds. The final cohort used for primary inference thus included n=119 SCA1 and n=126 CONT participants. Supplementary analyses were undertaken using the initial cohort for corroboration of the main results.

Demographic and clinical data for the final cohort are summarized in **Table 1**. The SCA1 and CONT cohorts were age- and sex-matched within each site, and in aggregate showed no significant differences in sex nor age. READISCA is considered as a single site for this purpose, despite data coming from multiple research centres, due to prospective harmonisation of scanning hardware and protocols.

**Table 1.** Summary of participant demographics and clinical characteristics in the final dataset.

| Site | SCA1 |  |  |  |  |  |  | CONT |  |  |
| --- | --- | --- | --- | --- | --- | --- | --- | --- | --- | --- |
|  | N | Sex, F (%) | Age | Ataxia Duration | CAG Repeats | SARA | Pre-Ataxic (%) | N | Sex, F (%) | Age |
| Campinas | 26 | 10 (38.5) | 44.6 (8.3) | 7.3 (6.4) | 44.3 (4.2) | 14.2 (6.5) | 1 (3.8) | 31 | 10 (32.3) | 45.5 (9.3) |
| Essen/Halle | 16 | 10 (62.5) | 47.4 (11.6) | 9.5 (4.2) | 46.3 (5.0) | 14.0 (5.8) | 1 (6.3) | 14 | 9 (64.3) | 46.9 (11.9) |
| Florence | 12 | 8 (66.7) | 46.2 (10.6) | 10.6 (6.1) | 55.0 (7.9) | 18.1 (12.6)<sup>a</sup> | 3 (25.0) | 12 | 8 (66.7) | 45.7 (13.7) |
| Milan | 16 | 8 (50.0) | 48.1 (12.8) | 5.2 (4.3) | 42.1 (3.9) | 6.7 (5.3) | 2 (12.5) | 18 | 12 (66.7) | 41.8 (9.8) |
| *Milan A* | 9 | 6 (66.7) | 55.9 (8.9) | 4.6 (3.8) | 40.7 (2.9) | 7.5 (6.2) | 1 (11.1) | 10 | 7 (70.0) | 47.5 (5.5) |
| *Milan B* | 7 | 2 (28.6) | 38.1 (9.9) | 6.2 (5.1) | 44.0 (4.4) | 5.7 (4.3) | 1 (14.3) | 8 | 5 (62.5) | 34.6 (9.5) |
| Minnesota | 15 | 9 (60.0) | 51.8 (10.2) | 10.9 (5.0) | 44.3 (2.8) | 8.5 (3.6) | 2 (13.3) | 20 | 8 (40.0) | 52.5 (16.4) |
| Paris | 15 | 6 (40.0) | 42.8 (15.3) | 6.6 (6.1) | 47.1 (6.6) | 10.7 (6.3) | 2 (13.3) | 15 | 6 (40.0) | 44.9 (12.6) |
| READISCA | 15 | 12 (80.0) | 42.1 (8.9) | 10.1 (7.6) | 43.5 (2.4) | 4.1 (3.5) | 6 (40.0) | 12 | 6 (50.0) | 41.3 (7.5) |
| Tübingen | 4 | 2 (50.0) | 27.0 (10.4) | --<sup>b</sup> | 47.5 (4.5) | 0.9 (1.2) | 4 (100) | 4 | 2 (50.0) | 27.8 (11.1) |
| **Total** | **119** | **65 (54.6)** | **45.4 (11.7)** | **8.2 (5.9)** | **45.1 (5.0)** | **10.6 (7.6)** | **21 (17.6)** | **126** | **61 (48.4)** | **45.2 (12.4)** |

Parentheses indicate standard deviation except where indicated. Age and ataxia duration are measured in years.
“Milan A” and “Milan B” denote subjects scanned with two different protocols. N: number of subjects; F: number of female subjects; SARA: Scale for the Assessment and Rating of Ataxia score. <sup>a</sup> Converted from equivalent ICARS score<sup>19</sup>; <sup>b</sup> Not applicable; all participants were pre-ataxic

### Image processing

All T_1_-weighted images were pre-processed using a previously described pipeline for the ENIGMA-Ataxia project^20^, which combines specialized software for analyzing the cerebellum and cerebrum.^21^ First, a cerebellar mask was derived for each image, either manually or using the ACAPULCO^22^ algorithm (v0.2.1). The automated masks were then visually inspected, with minor errors corrected manually and major errors excluded. Next, regional cerebellar GM volume was estimated using the Spatially Unbiased Infratentorial Toolbox (SUIT)^23^ v3.2 for SPM12^24^ v7771, a statistical parametric mapping suite built for MATLAB. The images were segmented into GM and WM partial volumes using the previously generated cerebellar mask; these volumes were then DARTEL normalized and resliced into SUIT space, with Jacobian modulation used to ensure that the intensity value assigned to each voxel in SUIT space was proportional to its original volume. The resulting cerebellar GM images were inspected for normalization errors, with particular attention given to outliers detected by calculating the spatial covariance of each image within the data set. If any processing errors occurred, these images were rejected, before finally being spatially smoothed using a Gaussian kernel with a full width at half-maximum (FWHM) of 3 mm.

Because SUIT is optimized for cerebellar GM, regional WM and cerebral GM volume were instead estimated using the Computational Anatomy Toolbox (CAT12)^25^ v12.5 for SPM12. The raw images were bias-field corrected, skull-stripped, then segmented into GM, WM, and cerebrospinal fluid (CSF) partial volumes, from which intracranial volume (ICV) could be calculated. The GM and WM partial volumes were DARTEL registered to the Montreal Neurological Institute (MNI) standard space, again using Jacobian modulation to ensure accurate volume encoding. The GM image was masked to remove the cerebellum to minimize undue influence on the cerebral results. Finally, both the GM and WM images were smoothed using a 5 mm FWHM Gaussian kernel.

### Statistical analysis

Demographic and clinical analyses were analyzed using R v4.4.2. The distribution of ages in each group was checked for normality using the Shapiro-Wilk test: if either group was non-normally distributed, the Wilcoxon ranked-sum test was used to determine if groupwise differences were significant; otherwise, Student’s T-test was used. Sex was compared between groups using Fisher’s exact test.

Image analyses were performed using SPM12 in MATLAB. In all cases, heteroscedasticity was assumed and corrected for, with final voxel-level inferences estimated using random field theory to account for the large number of multiple comparisons endemic to imaging analysis.^26^ Family-wise error (FWE) correction was set at the *α* = 0.05 level, with only significant clusters of *k* > 100 voxels being reported. Statistical maps showing significant results were then converted to Cohen’s *d* (for groupwise analyses) or Pearson’s *r* (for correlation analyses) to describe effect size.^27,28^

### Groupwise analysis for disease effects

Between-groups comparisons of the final SCA1 and CONT cohorts were performed using a general linear model (GLM) in SPM12 with group (2 levels, i.e. SCA1 and CONT) and site (8 levels) as factors, and ICV and age as covariates. Of these, only group is a comparison of interest; the others serve as nuisance variables. This analysis was repeated in the initial dataset using one factor (group, 2 levels) and two nuisance covariates (ICV, age).

For interpretive purposes, the voxel-based results were mapped onto the following anatomical atlases: the Harvard-Oxford cortical and subcortical GM atlases and Johns Hopkins University cortical WM atlas, all included with FSL^29^; the SUIT cerebellar GM and dentate nucleus atlases^30,31^; the van Baarsen cerebellar WM atlas^32^; and the FreeSurfer brainstem atlas.^33^ Significant results in cerebellar GM were additionally mapped onto the multi-domain task-based (MDTB) parcellation developed by King *et al*.^*16*^ to provide structure-function insights.

### Clinical correlations for SCA1 participants

In the final SCA1 cohort, the effects of healthy aging^34^ and site-specific confounds^35^ were estimated and adjusted for prior to undertaking clinical correlations using previously reported methods.^36^ These age- and site-adjusted SCA1 images were assessed for linear relationships between volume and ataxia duration, SARA score, or the long CAG triplet repeat length on *ATNX1*. In pre-ataxic participants for whom both the long and short CAG triplet counts were available, time to onset was estimated using an established formula^37^ and recorded as a negative ataxia duration. For each correlation, ICV was included as a nuisance variable to account for subject head size.

For visualisation, whole-brain effect maps and scatterplots of volume vs. ataxia duration from selected regions of interest (ROI) are presented. ROI volume was z-normalized based on the CONT distribution, controlling for age, as previously described.^38^ ROIs were selected to represent areas of high effect size within each of the three components of the brain analyzed, *i*.*e*., WM, cerebellar GM, and cerebral GM.

### Ataxia severity stratification

Cross-sectional analyses were performed on subsets of the SCA1 participant data to track disease evolution. The data were subdivided into five groups – pre-ataxic (SARA < 3)^39^ and four quartiles of ataxia-manifest individuals (quartile separation values: SARA = 8, 11, 17.5) – which were then compared to the full CONT cohort. The age- and site-corrected final SCA1 and CONT dataset was used, and ICV was included as a nuisance variable.

A similar cross-sectional analysis was undertaken to qualitatively examine the interaction between CAG repeat length and ataxia duration. The final dataset was subdivided into four bins using the median split of each variable. Again, SCA1 subject data in each case were compared to the CONT cohort following age and site correction, with ICV as a nuisance covariate. Finally, a regression model was calculated for each ROI previously highlighted for clinical correlations, considering main effects of CAG repeat length and ataxia duration and the interaction of CAG × Duration, with age and ICV as nuisance covariates.

### Cerebrocerebellar covariance

To better understand the relationship between disease effects in the cerebrum and cerebellum, we performed a cerebrocerebellar structural covariance analysis, using the following linear model:

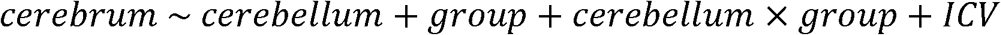

Where *cerebrum* and *cerebellum* represent the GM, WM, or GM+WM volumes in those respective regions of the brain. For this analysis, the CAT12-derived WM was cropped to the cerebrum, while the SUIT-derived cerebellar WM (including the brainstem) that is normally discarded was retained. To visualize these relationships, partial regression plots were created following correction for ICV.

Because this analysis examined group differences in the extent to which cerebellar atrophy is linked to cerebral atrophy, the *cerebellum* × *group* interaction was the effect of interest. A stronger correlation in the SCA1 group than in CONT would indicate that atrophy in the cerebellum is coupled to changes in cerebrum, while a weaker correlation in SCA1 relative to CONT would suggest a relative independence of the cerebellar and cerebral volumetric changes.

## Results

### Groupwise volumetrics in SCA1 and control participants

Group differences between SCA1 and CONT in the final cohort are presented in **Figure 1**, and in the initial cohort in Supplementary Figure S1. The results in both cohorts were highly similar. The greatest effect sizes (*d* > 1.0) were found bilaterally in the cerebellar WM and brainstem (**Figure 1A**), including the dentate region, medial lemnisci, pons and pontine crossing tract, corticospinal tracts, cerebral and cerebellar peduncles, medulla, and midbrain. Moderate to strong atrophic effects (0.5 < *d* < 1.0) were observed in the bilateral internal capsule and superior corona radiata, as well as the entirety of the cerebellar GM (**Figure 1B**), peaking in bilateral lobules I-IV and VI, vermis IX, and left lobule V. Moderate atrophy was also observed in the bilateral caudate and superior putamen and right superior frontal gyrus in the cerebral GM (**Figure 1C**). Repeating the analysis on the larger initial dataset returned a few superficial differences from the main results in the cerebral GM (**Supplementary Figure S1A, B**) and some slight decreases in effect size in the cerebral WM and cerebellar GM (**Supplementary Figure S1C, D**).

**Figure 1.**
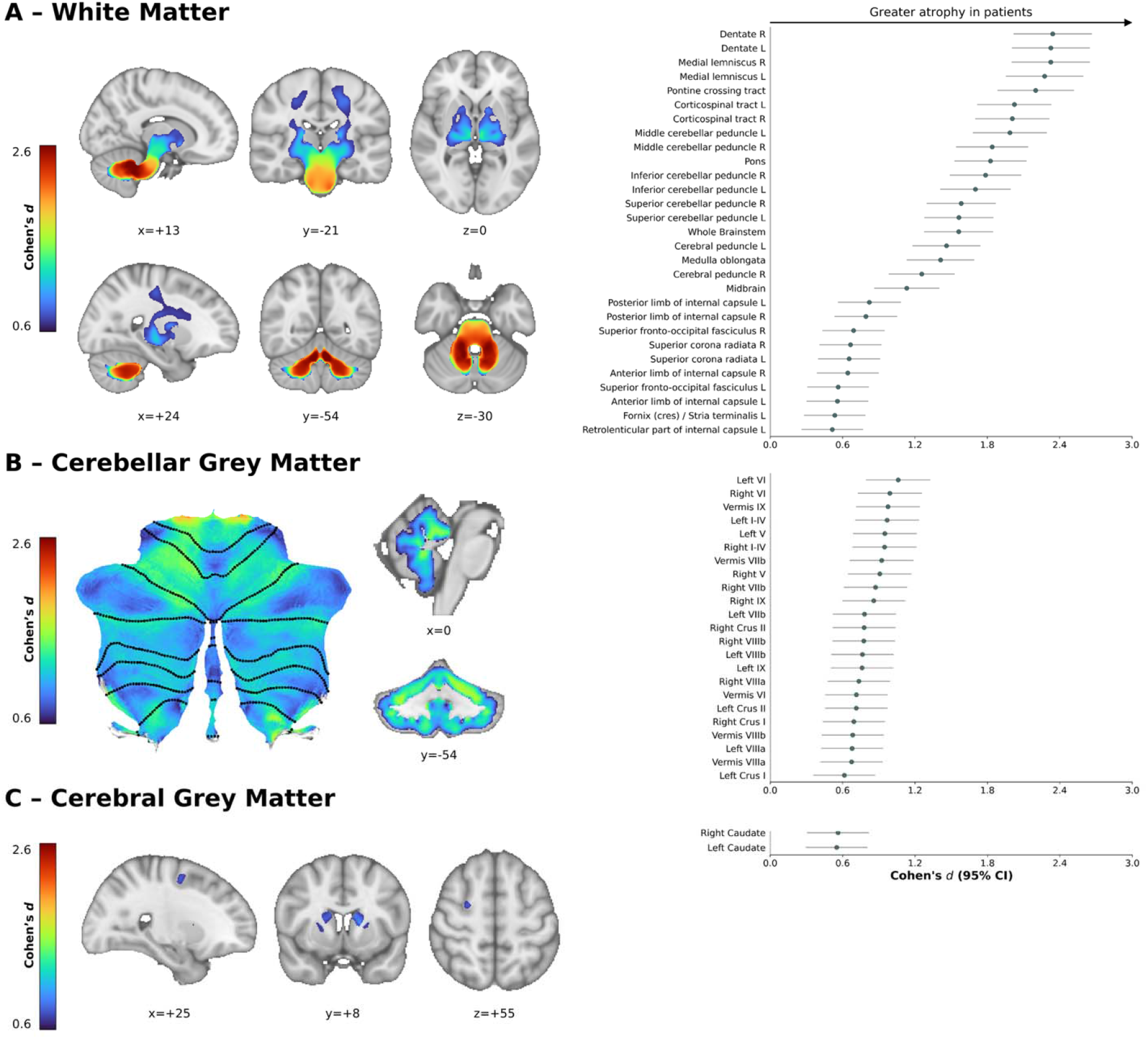
Regions of significantly lower volume (voxel-level FWE-corrected p < 0.05) in participants with SCA1 relative to CONT in (A) whole-brain white matter, (B) cerebellar grey matter, and (C) cerebral grey matter. Left: representative slices or cerebellar flatmaps illustrating the areas of significant atrophy in SCA1 subjects. Right: forest plots illustrating regional effects (Cohen’s d > 0.5, i.e., a moderate effect size); error bars represent the 95% confidence interval (CI). Slice coordinates are in Montreal Neurological Institute (MNI) space.

### Network representation of groupwise cerebellar GM atrophy

The SCA1 atrophy profile within the cerebellar GM was subsequently mapped onto the MDTB functional atlas (**Figure 2**). From this, we observed that the greatest atrophy occurred in MDTB Regions 5-7, representing attention, executive function, and language/emotional processing. Slightly lower atrophic effect size was seen in regions responsible for motor planning (Regions 1 and 2) and language/verbal processing (Regions 8 and 9). Weaker effects were seen in the remaining regions, which have been ascribed to visuospatial and cognitive tasks, visual working memory, and saccades. Note that the functions ascribed to these regions are representative, not exhaustive.

**Figure 2.**
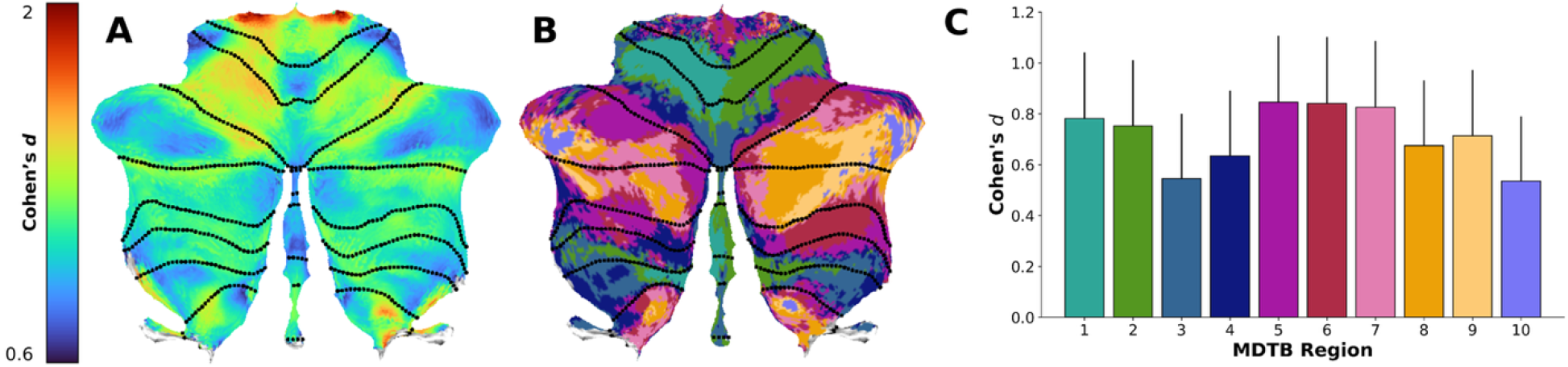
Mapping of structural changes to functional networks in the cerebellum. (A) Cerebellar flatmap of the voxel-level effect size of volume differences in participants with SCA1 vs. CONT in cerebellar grey matter, rescaled from Figure 1B. (B) Cerebellar flatmap of the multi-domain task battery (MDTB) functional atlas from King et al.^16^. (C) A bar chart of the effect sizes by MDTB region; bar height is the mean effect size within each region, while error bars are the positive 95% CI. The colours of the bars in Panel C match the colours of the regions in Panel B.

### Clinical correlations in SCA1 participants

Voxelwise correlations were calculated between volume and both ataxia severity and duration (**Figure 3**). The strongest correlations were observed between SARA score and volume in the cerebellar WM, with correlation coefficients reaching |*r*| = 0.80. Slightly lower correlation coefficients (0.60 < |*r*| < 0.80) were observed between SARA and volume in the pons and portions of the cerebellar GM, specifically bilateral lobule VIIb and left lobules VIIIa/b and XI. Correlation coefficients in this range were also observed between ataxia duration and cerebellar WM and pontine volumes. The weakest significant correlations (0.40 < |*r*| < 0.60) occurred between SARA and volume in cerebral WM and GM, specifically in the bilateral internal capsule and right putamen, and between ataxia duration and scattered portions of the cerebellar GM. CAG repeat length on the long allele also correlated with volume in bilateral lobule VI (**Supplementary Figure S2**).

**Figure 3.**
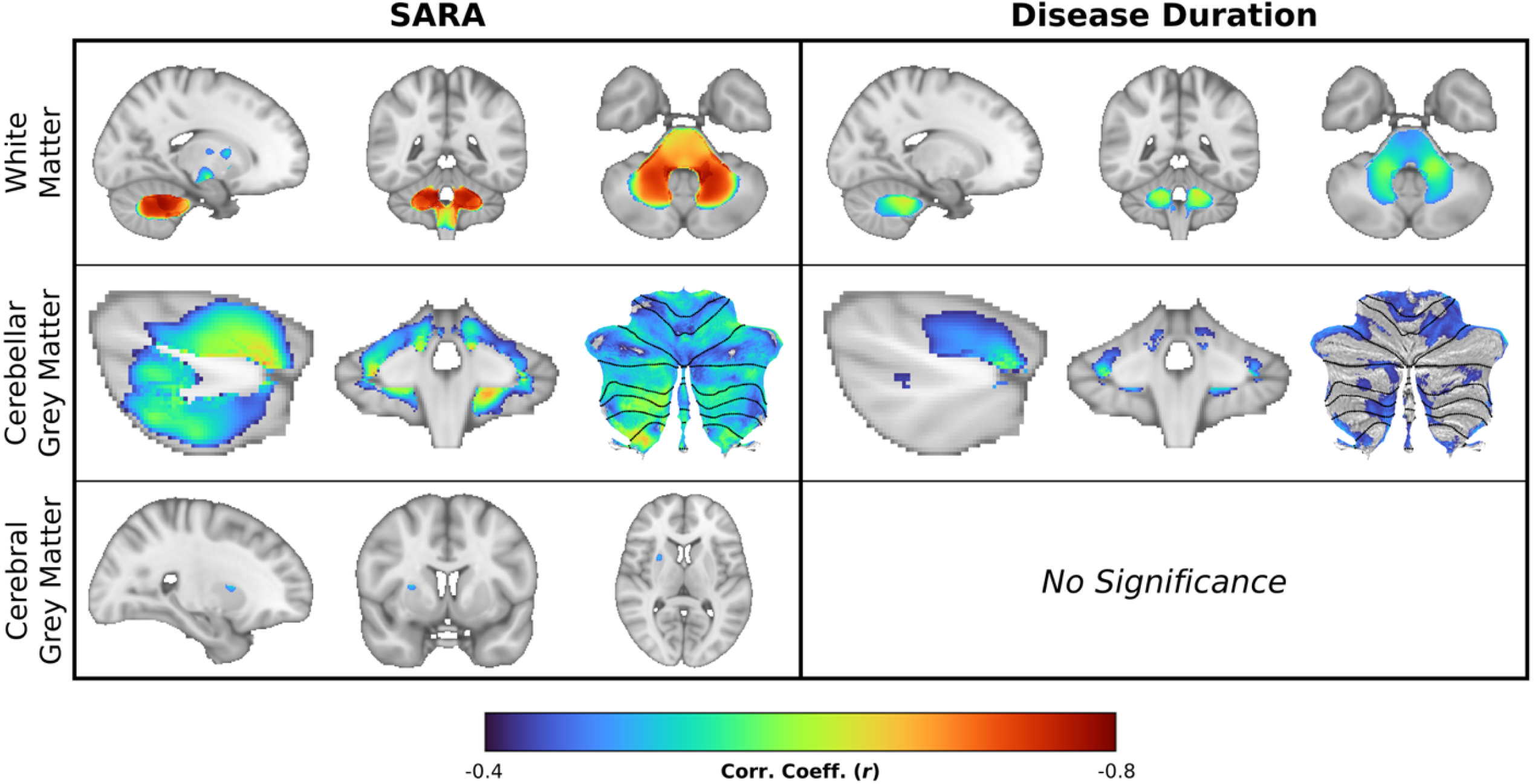
Correlations between volume and ataxia severity (SARA score, left) and ataxia duration (right) in SCA1 participants (voxel-level FWE-corrected p < 0.05). Top row: whole-brain white matter at MNI coordinates (-17,-43,-38). Middle row: cerebellar grey matter at MNI coordinates: x = +34 and y = -42 with cerebellar flatmap. Bottom row: cerebral grey matter at MNI coordinates (+25,+6,+8).

Representative scatterplots of these relationships are presented in **Figure 4**. The WM regions chosen – the bilateral dentate regions and the pons – both showed very strong correlations, with a clear progression from pre-ataxic to ataxic disease stages. The cerebellar GM regions – bilateral lobules I-IV and VI – both showed moderate effects with clear downward trends. In the two selected striatal regions, the putamen volume correlated modestly but significantly with ataxia duration, while the caudate volume did not correlate significantly, despite the caudate having a stronger overall groupwise effect size and a larger area of significant voxelwise atrophy (**Figure 1C**).

**Figure 4.**
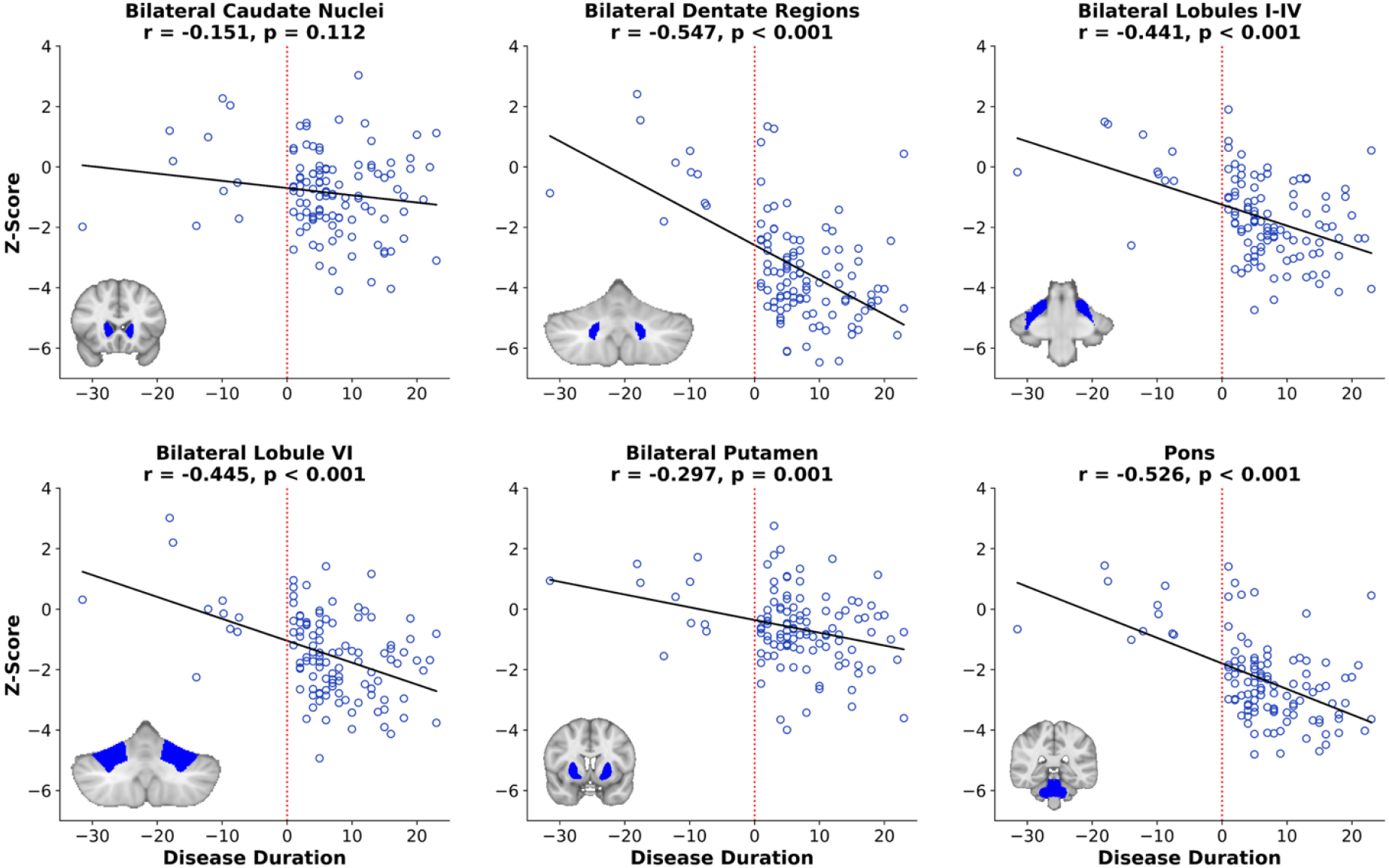
Individual variability within selected regions of interest (ROIs). ROI volume in SCA1 participants is converted to a Z-score within the distribution of CONT patients, then plotted against ataxia duration. Solid line: linear fit between Z and disease duration; dashed line: duration = 0 line, denoting the separation between pre-ataxic and ataxic SCA1 participants. At the bottom left corner of each plot is an anatomical reference for each ROI’s position in the coronal plane.

### SCA1 severity stratification

To further characterize the evolution of atrophy over the course of SCA1, the clinical cohort was stratified by SARA score into pre-ataxic and four quartiles of ataxia severity and individually compared to a common control cohort (**Figure 5**). As a general trend, atrophy becomes more severe and widespread with greater disease severity, though the spread of atrophy is comparatively limited after the second quartile. In the WM, atrophy begins in the cerebellum and medulla in the pre-ataxic cohort before spreading to the pons in the first quartile, then outward along the corticospinal tract through the internal capsule and superior corona radiata before terminating near the sensorimotor cortex beginning in the second quartile. Greater magnitude of atrophy, but limited spread, occurs beyond this point.

**Figure 5.**
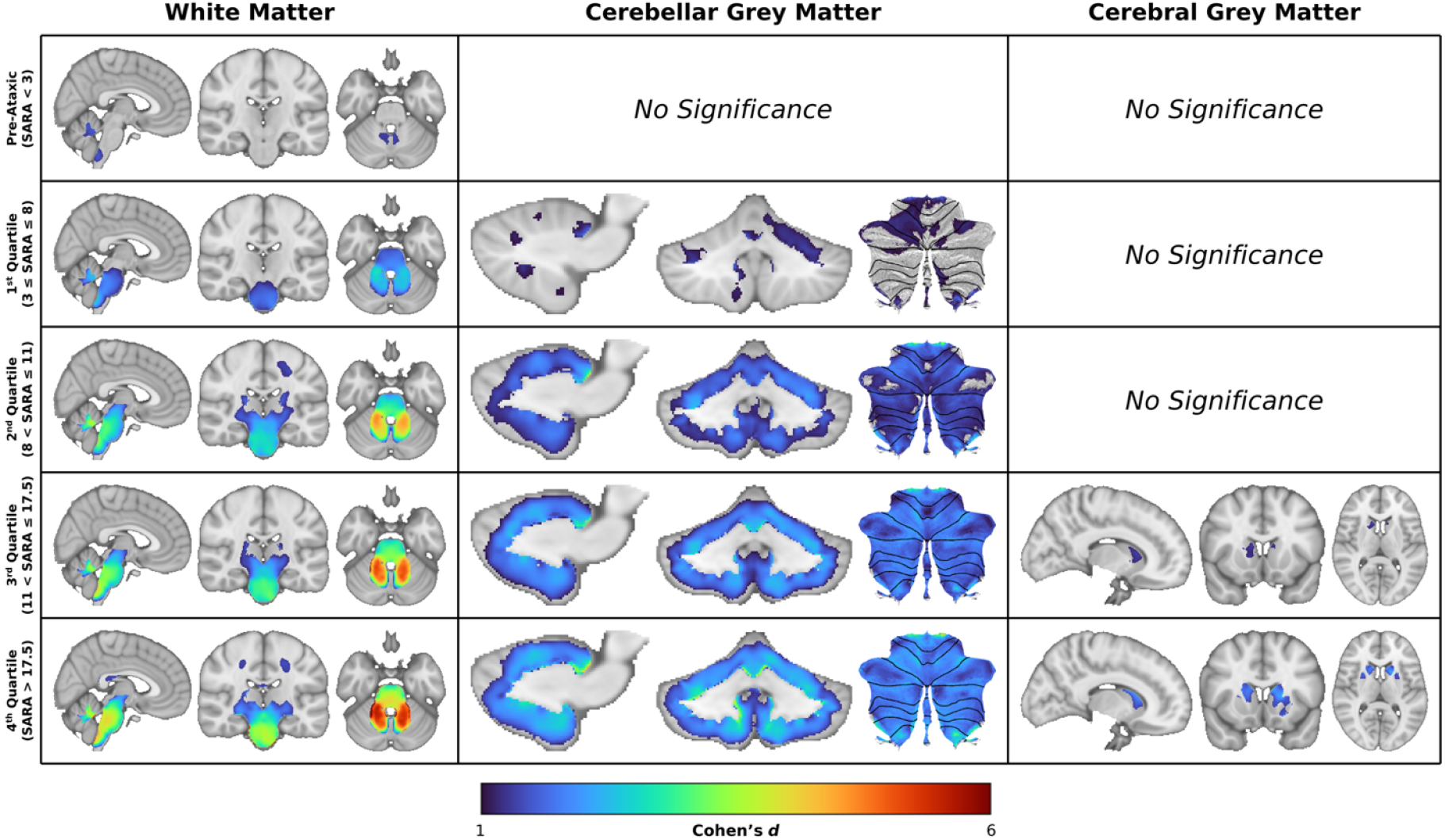
Stratification of the SCA1 cohort based on SARA score. Voxel-level effect size maps (FWE-corrected p < 0.05) of each SCA1 subgroup relative to CONT. From top to bottom: Pre-ataxic (SARA < 3; n = 21), 1^st^ Quartile (3 ≤ SARA ≤ 8; n = 26), 2^nd^ Quartile (8 < SARA ≤ 11; n = 22), 3^rd^ Quartile (11 < SARA ≤ 17.5; n = 27), 4^th^ Quartile (SARA > 17.5; n = 20). All images show representative slices or flatmaps of the affected tissue type: white matter (left; MNI coordinates +4,-21,-29), cerebellar grey matter (centre; MNI x = +14 and y = -52 plus flatmap), and cerebral grey matter (right; MNI coordinates +12,+13,+9).

In the cerebellar GM, atrophy is non-significant in the pre-ataxic stage and largely limited to the anterior lobe (lobules I-V), lobule VI, and lobule IX in the first quartile. From the second quartile onward, there is significant atrophy across the entire cerebellum, which increases in magnitude in the third and fourth quartiles. Finally, in the cerebral GM (right column), no differences are significant until the third quartile, at which point atrophy in the right caudate and small parts of the left become significant; this progresses to encompass the full bilateral caudate and small parts of the bilateral putamen in the fourth quartile.

The significance threshold in this comparison requires an effect size more than Cohen’s d > 1.0, representing a large effect size. To mitigate the potential for Type II error in our data interpretations, the stratified data were also re-plotted at a reduced threshold of *d* > 0.5 and examined qualitatively (**Supplementary Figure S3**). As expected, this data depiction shows the smaller effects that precede detection in the statistically robust main analysis, *e*.*g*., pontine involvement in the pre-ataxic stage, and striatal effects in early symptomatic disease. This also suggests a broader, but more subtle profile of cerebral GM involvement that is not seen in the main analysis. However, importantly, the pattern of these effects is not consistent across the stratified subgroups, and there is no apparent pattern of worsening or expanding cerebral cortical involvement with disease evolution. These subthreshold cerebral GM effects therefore potentially reflect important aspects of individual disease expression and inter-individual heterogeneity, but not common core features of SCA1.

### SCA1 CAG x duration stratification

Stratification of the sample was then undertaken using CAG repeat length and ataxia duration in combination (**Figure 6**; subsample demographic and clinical characteristics reported in **Supplementary Table S2**). Regression analyses using the same ROIs as in the overall disease duration correlation assessments (see Figure 4) indicated significant CAG-by-duration interaction effects in the cerebellar GM (ROIs: Lobule I-IV and Lobule VI). As visualised in Figure 6A, and plotted in Figure 6B, this result was driven by less atrophy in people with fewer CAG repeats early in the disease course, but comparable atrophy in later disease. A main effect of CAG repeat length, but not a significant interaction with disease duration, was also evident in the cerebellar WM (dentate region ROI) and pons. This result was driven by greater WM atrophy in people with greater CAG repeat length regardless of duration of ataxia (Figure 6A, B). No significant main or interaction effects were evident for the striatal ROIs (caudate and putamen).

**Figure 6.**
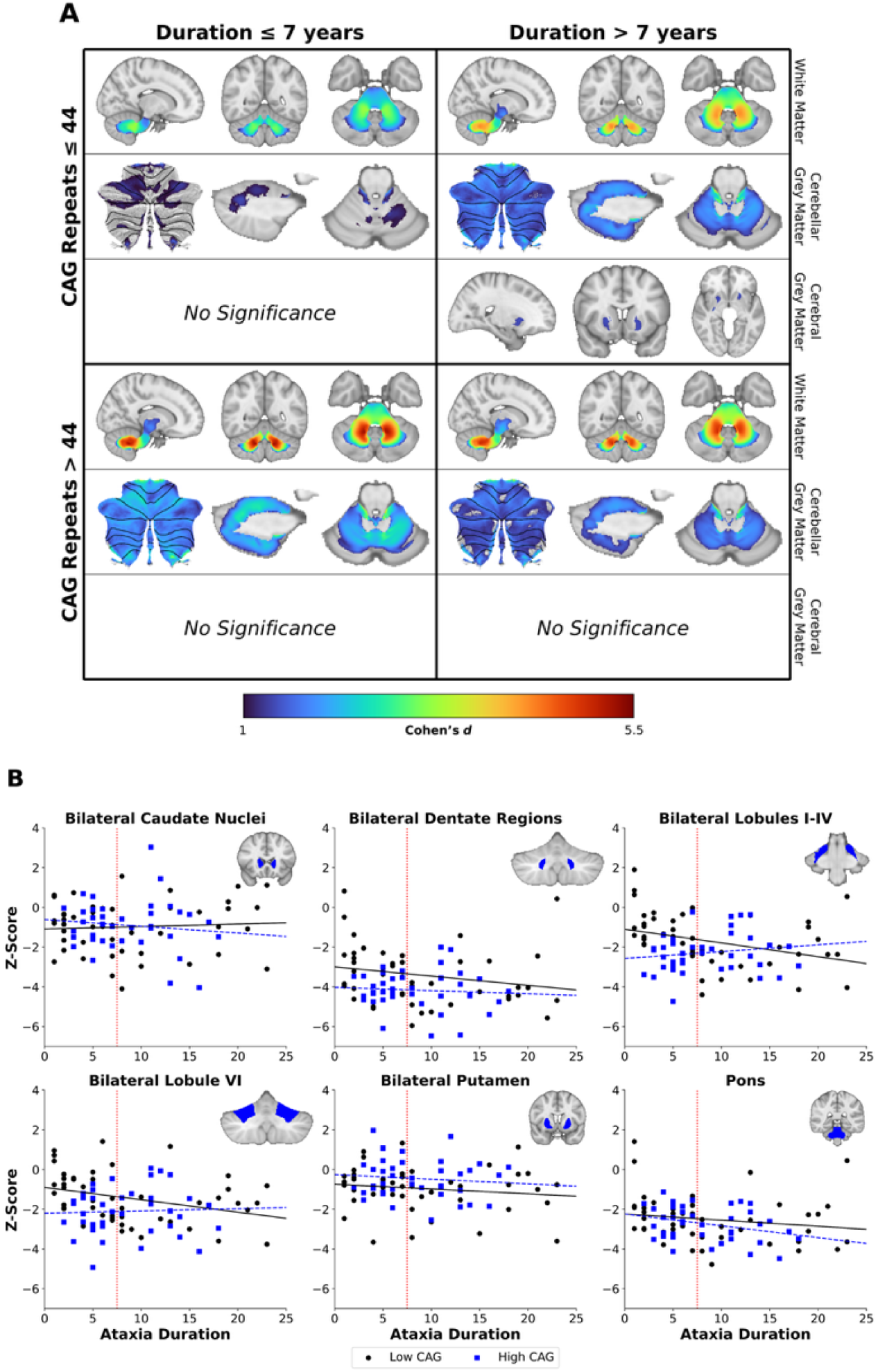
Relationships between ataxia duration and CAG repeat length. (A) The cohort was divided into four subgroups based on the median value of each variable: low duration, low CAG (top left; n = 27); high duration, low CAG (top right; n = 21); low duration, high CAG (bottom left; n = 21); high duration, high CAG (bottom right; n = 19). Rows within each quadrant are, from top to bottom: whole-brain white matter (MNI coordinates: +14,-52,-36); cerebellar grey matter (flatmap, MNI x = -17, and MNI z = -24); and cerebral grey matter (MNI coordinates +22,+12,-6). (B) Scatterplots of normalized ROI volumes as previously shown in Figure 4, stratified based on CAG repeat count (“Low” represents CAG ≤ 44; “High” represents CAG > 44); the red dotted line represents the median of ataxia duration. Black circles and solid lines represent low CAG repeat count; blue squares and dashed lines represent high CAG repeats.

**Figure 7.**
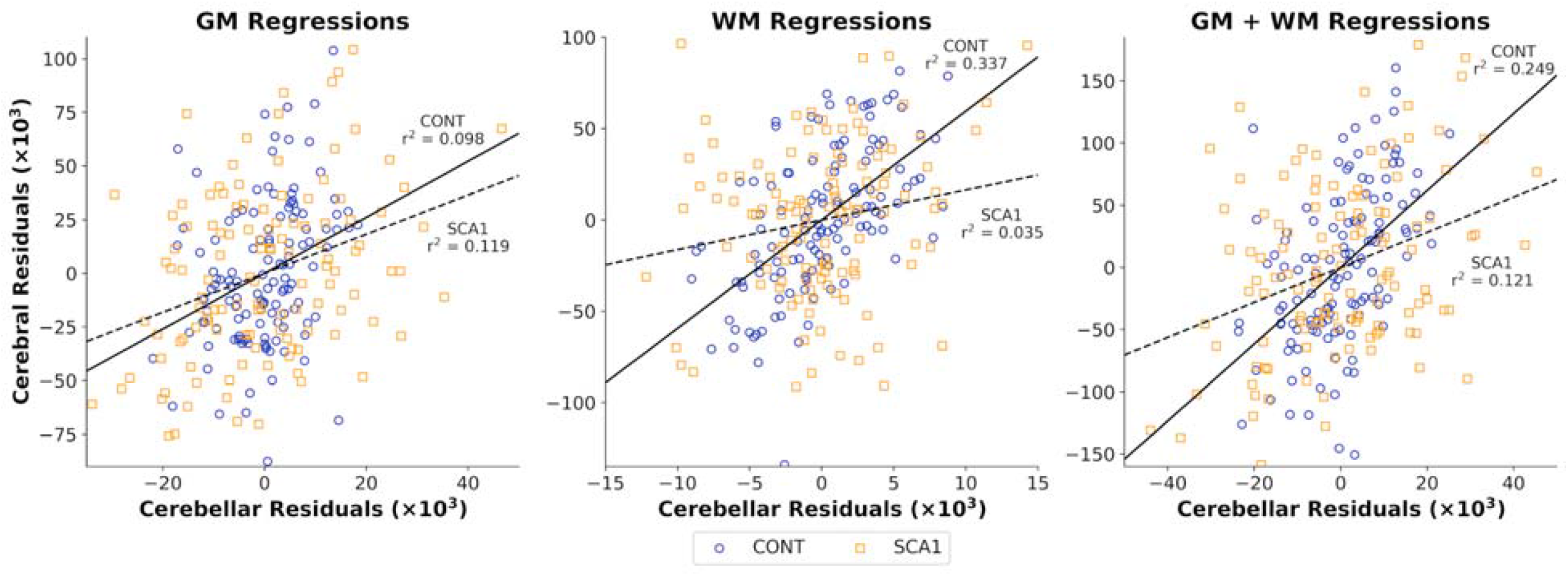
Cerebrocerebellar spatial covariance partial regression plots. Axes represent the cerebral (vertical) and cerebellar (horizontal) volumes residuals after correcting for ICV in grey matter (left), white matter (center), and grey+white matter (right). Blue circles: residual volumes in CONT subjects; orange squares: residual volumes in SCA1 subjects.

### Cerebrocerebellar covariance

A significant positive correlation was observed between cerebellum volume and cerebral volume of WM, GM, and WM+GM in both CONT and SCA1 groups (SCA1 WM: *p* = 0.041; all others: *p* < 0.001). Significant group differences in the strength of these effects (i.e., *cerebellum* × *group* interactions) were evident in the WM (*p* = 0.004) and GM+WM (*p* = 0.007), but not GM alone (*p* = 0.14); see **Supplementary Table S3** for full statistics reporting. Partial regression plots indicated that these group differences resulted from a reduced strength of cerebrocerebellar covariance in the SCA1 cohort relative to the CONT cohort.

## Discussion

In this multi-site study aggregating a relatively large SCA1 participant cohort, we demonstrate that SCA1 is characterised by robust volume differences relative to healthy individuals in the cerebellum, brainstem, and basal ganglia, as well as the tracts connecting these regions to each other and to the somatomotor regions of the cerebral cortex. Ataxia severity correlates strongly with atrophy in the cerebellar WM and brainstem, and to a lesser extent with regional cerebellar GM. Mapping SCA1 atrophy onto a functional atlas of the cerebellum revealed preferential involvement of areas implicated in motor generation, attention, and executive function. Cross-sectional stratification across different levels of ataxia severity showed a spatial expansion of atrophy over the early course of the disease, while the more severely ataxic groups were predominately defined by progressive atrophy within already affected regions. CAG repeat length modified the relationship between ataxia duration and atrophy magnitude in the cerebellar cortex. Finally, we observed that the association between cerebral and cerebellar WM volumes was reduced in individuals with SCA1 relative to controls.

### Atrophy in SCA1 is predominant in Cerebellar WM and Pons

The atrophy profile reported here is generally consistent with previous reports in smaller cohorts.^3,4,13,14,5–12^ Robust atrophy was observed through the entirety of the cerebellar WM and brainstem – especially the pons – as well as WM tracts including the cerebral and cerebellar peduncles, medial lemnisci, corticospinal tracts, and pontine crossing tract. Moderate to strong effects were similarly seen throughout the internal capsule and the superior corona radiata. Our GM findings similarly agree with the majority of published literature, with extensive atrophy throughout the cerebellar GM as well as the bilateral caudate and putamen.

Contrary to some other studies showing a bias in the atrophy toward the right side of the brain^3,14,15^, we did not find evidence of lateralisation. Additionally, several studies have reported damage to the corpus callosum^3,6,8,9^, which we replicated only in the most severely affected disease subgroup in the current analysis. Similarly, previous studies have reported cortical thinning or volume loss in the cerebral cortex, particularly in later disease stages, which we only observed at sub-threshold significance levels.^6,8^ These distinctions may reflect the impact of individual variability on previous outcomes in relatively small samples. Equally, our scatterplots of regional volumes relative to ataxia duration demonstrate a marked spread of individual outcomes, even in core disease regions (i.e., pons). Taken together, it is likely that cerebral atrophy may represent an important aspect of inter-individual variability in SCA1 manifestation, but it is not a universal element of the disease signature.

In line with previous reports, the strength of correlations with ataxia severity mirrored the spatial profile of atrophy^3,4,8,9,11,12,14^. Large correlations with magnitudes in the range of 0.6-0.8 were observed in regions of the cerebellar WM, pons, and select areas of the cerebellar cortex. These findings are in line with recent work showing similarly strong cross-sectional correlations with SARA^9,11^, and longitudinal sensitivity of pons volume and MCP diffusion measures in SCA1 cohorts.^7,11,12,15^ Our full-brain voxelwise analyses replicate and add greater spatial information to the profile of clinico-anatomical relationships in SCA1.

Finally, mapping cerebellar atrophy onto the functional atlas of King *et al*.^*16*^ showed preferential atrophy in both motor and cognitive regions in individuals with SCA1. This is consistent with previous observations we have made in SCA2 and SCA3^40,41^, and although the present study does not include cognitive data, it is in line with growing body of work showing high rates of CCAS in people with polyglutamine SCAs.^6,8,42,43^ Future work pairing of cognitive testing with imaging is necessary to definitively link CCAS with a particular profile of cerebellar degeneration, and to establish the prognostic value of imaging in predicting those at greatest risk of poor cognitive outcomes.^42^

### Stratified analysis of SCA1 atrophy

In this study, we have stratified SCA1 participants by ataxia severity to better understand its evolution. One previous study^6^ similarly separated SCA1 subjects by ataxia duration and produced findings with some similarities to ours: the group with the lowest ataxia duration (< 5 years) had significant WM atrophy only in the cerebellar peduncles and lower brainstem, which then spread to the remainder of the brainstem and beyond. In our data, this is comparable to the pattern of progression observed from the pre-ataxic subjects through the first two quartiles of ataxia severity, although we also show increasing involvement of the cerebellar cortex over this period. As noted above, we do not find strong evidence for the involvement of the cerebral cortex in SCA1, even at later disease stages.

While SARA staging is not equivalent to a longitudinal study, stratified cross-sectional analysis across the disease continuum serves as a useful proxy to understand the full disease course: multi-site, retrospective data pooling provides sample sizes large enough to undertake feasible and reliable stratified analyses. Longitudinal follow-up across the full multi-decade disease course currently remains infeasible at scale, and most longitudinal studies to date have focussed on 1–2 year periods for the purposes of biomarker discovery for use in clinical trials.^5,7,8,11,12,14,15^ However, our findings here agree with trends observed in longitudinal studies, including significant progressive atrophy in the brainstem, especially the pons, the cerebellar GM and WM, and the corticospinal tract and cerebellar peduncles.

Our examination of the interplay between ataxia duration and CAG repeat length on brain volume is consistent with the known clinical relationship between these measures, at least early in the disease course. CAG repeat length is associated with both earlier onset and more rapid progression of ataxia symptoms in SCA1.^2,37^ Here we show that a shorter CAG repeat length is associated with less cerebellar GM atrophy and lower average SARA scores, consistent with a milder disease presentation, but only in the early phases of the disease. Beyond the median of ataxia duration in our sample, differences in atrophy magnitude or SARA score were no longer evident. It is possible that these observations may be a function of floor effects, whereby volume loss reaches a maximum earlier in the disease course in people with a greater CAG repeat length.

### Cerebrocerebellar structural analysis

Finally, we examined the relationship between cerebral and cerebellar volumes in each group across GM, WM, and the sum of the two using a structural covariance analysis. Our results illustrate that the normal covariance between the cerebellar and cerebral WM and total volumes are significantly reduced in SCA1. This finding is consistent with but greater in magnitude than our previous report in people with SCA3.^41^ Attenuation of this relationship in individuals with SCA1 suggests that volume loss in the cerebellum occurs relatively independent of cerebral changes. This result adds further weight to the conclusion above that the consistent profile of cerebellar volume loss in people with SCA1 is not mirrored by a consistent cerebral anatomical pathology.

### Limitations

While our work brings new findings to the field, several limitations are notable. First, our data are restricted to structural T_1_-weighted images. In the future, it will be important to incorporate other modalities into our understanding of SCA1, such as diffusion or functional, as both can provide unique and useful information. Second, while stratification and correlation based on ataxia severity and duration provides insights into the evolution of SCA1, this remains a cross-sectional analysis of different individuals at different disease stages. Longitudinal data is necessary to investigate and validate individual-level brain atrophy. Finally, the retrospective nature of our study design limits the availability of clinical and behavioural measures across all contributing sites. In particular, cognitive and affective data are not available. Given the prevalence of CCAS in polyglutamine SCAs^6,8,42,43^, it would be useful to correlate measures of cognitive decline and/or emotional dysfunction with changes in brain volume to corroborate our findings of overlap between atrophy and cognitive regions of the cerebellar functional atlas.

## Conclusions

This study, using the largest multi-site sample of SCA1 participants and matched healthy controls to date, provides a robust and comprehensive profile of neurodegeneration in SCA1 carriers. Our results show a snapshot of a disease that spreads from the WM of the cerebellum and brainstem to the cerebellar GM, followed by cortical white matter underlying the somatomotor and sensory cortices. Structural covariance analysis shows a decoupling of cerebral and cerebellar volumes in WM in SCA1 participants, in line with strong infratentorial atrophy and limited evidence of core supratentorial involvement. These findings expand our knowledge of SCA1 and provide a number of fruitful avenues for future investigation.

## Data Availability

All data produced in the present study are available upon reasonable request to the authors

## Supplementary Figures

**Supplementary Figure S1.**
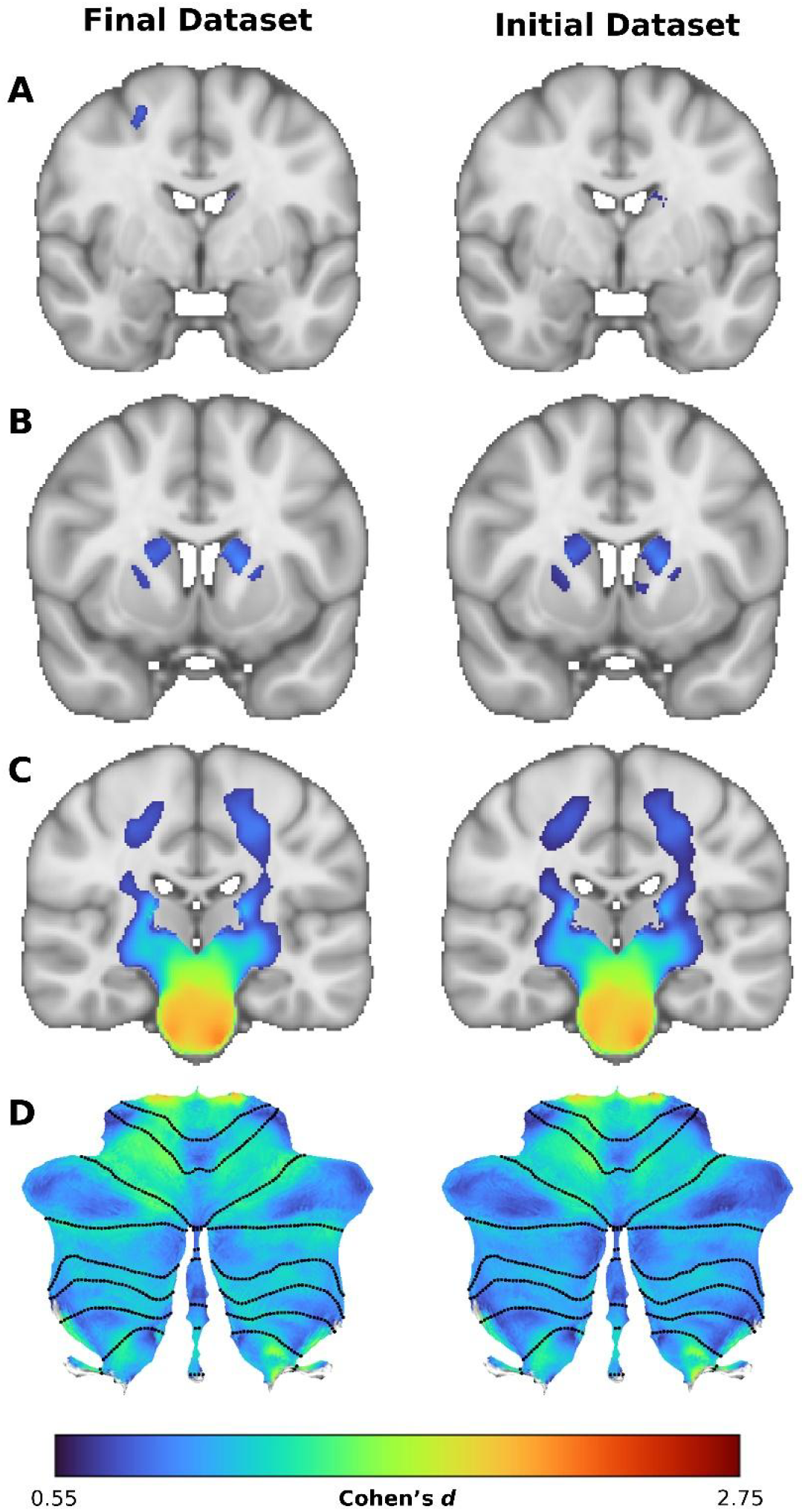
Images showing the differences between findings from the final dataset as reported in the main Results (left) and the larger, initial dataset without site correction (right). (A) Cerebral grey matter at y = -3, showing the loss of a significant region in the right superior frontal gyrus. (B) Cerebral grey matter at y = +8, showing a slight increase in the size of the significant regions of the bilateral putamen and caudate nuclei. (C) White matter at y = -21, reflecting a slight decrease in effect size. (D) Cerebellar grey matter flatmap, reflecting a slight decrease in effect size.

**Supplementary Figure S2.**
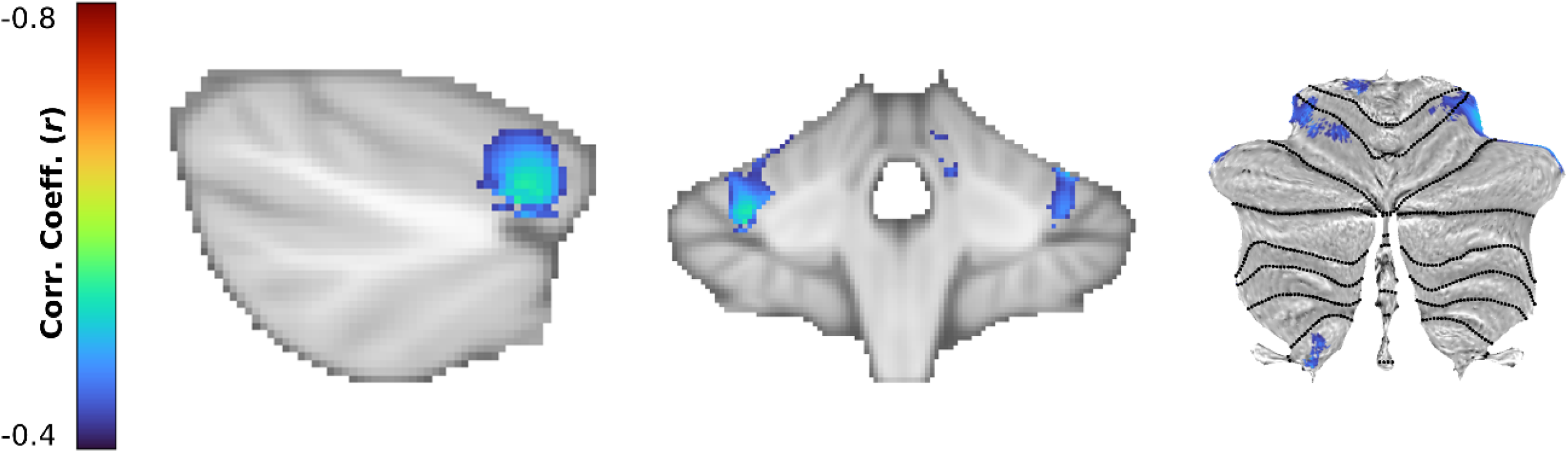
Correlation between cerebellar volume and CAG repeat length in SCA1 participants, taken at MNI coordinates x = +34 and y = -41 with cerebellar flatmap.

**Supplementary Figure S3.**
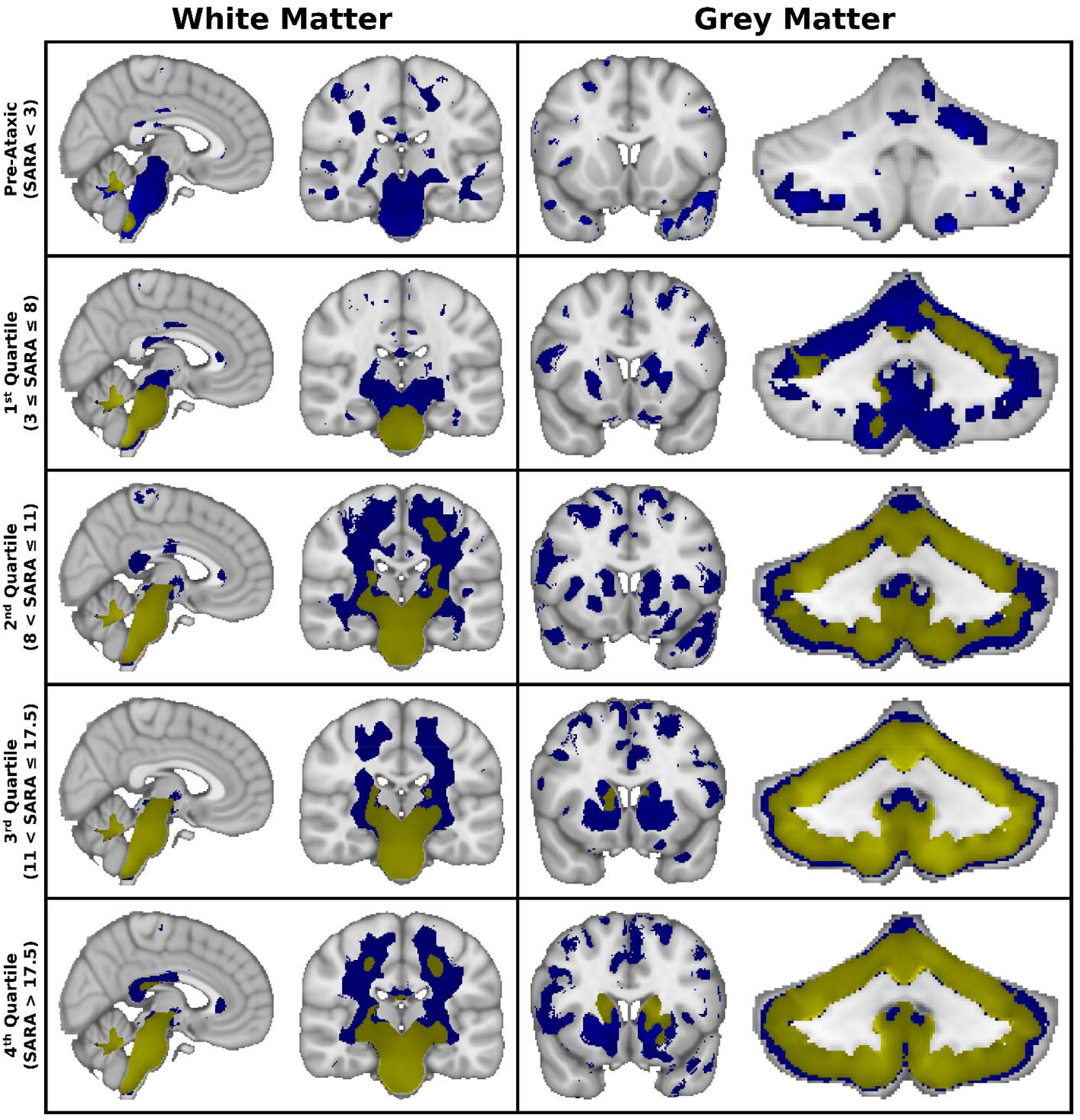
Selected views of non-significant trends in white matter (left) and grey matter (right) in SCA participants compared to controls, stratified by ataxia severity (SARA score). From top to bottom: Pre-Symptomatic (SARA < 3; n = 21), 1^st^ Quartile (3 ≤ SARA ≤ 8; n = 26), 2^nd^ Quartile (8 < SARA ≤ 11; n = 22), 3^rd^ Quartile (11 < SARA ≤ 17.5; n = 27), 4^th^ Quartile (SARA > 17.5; n = 20). Blue data represents Cohen’s d > 0.5, while yellow represents FWE-corrected p < 0.05, identical to the main analysis. MNI coordinates: x = +4 and y = -21 (white matter), y = +13 (cerebral grey matter, third from left), and y = -52 (cerebellar grey matter, rightmost)

## Supplementary Tables

**Supplementary Table S1.**
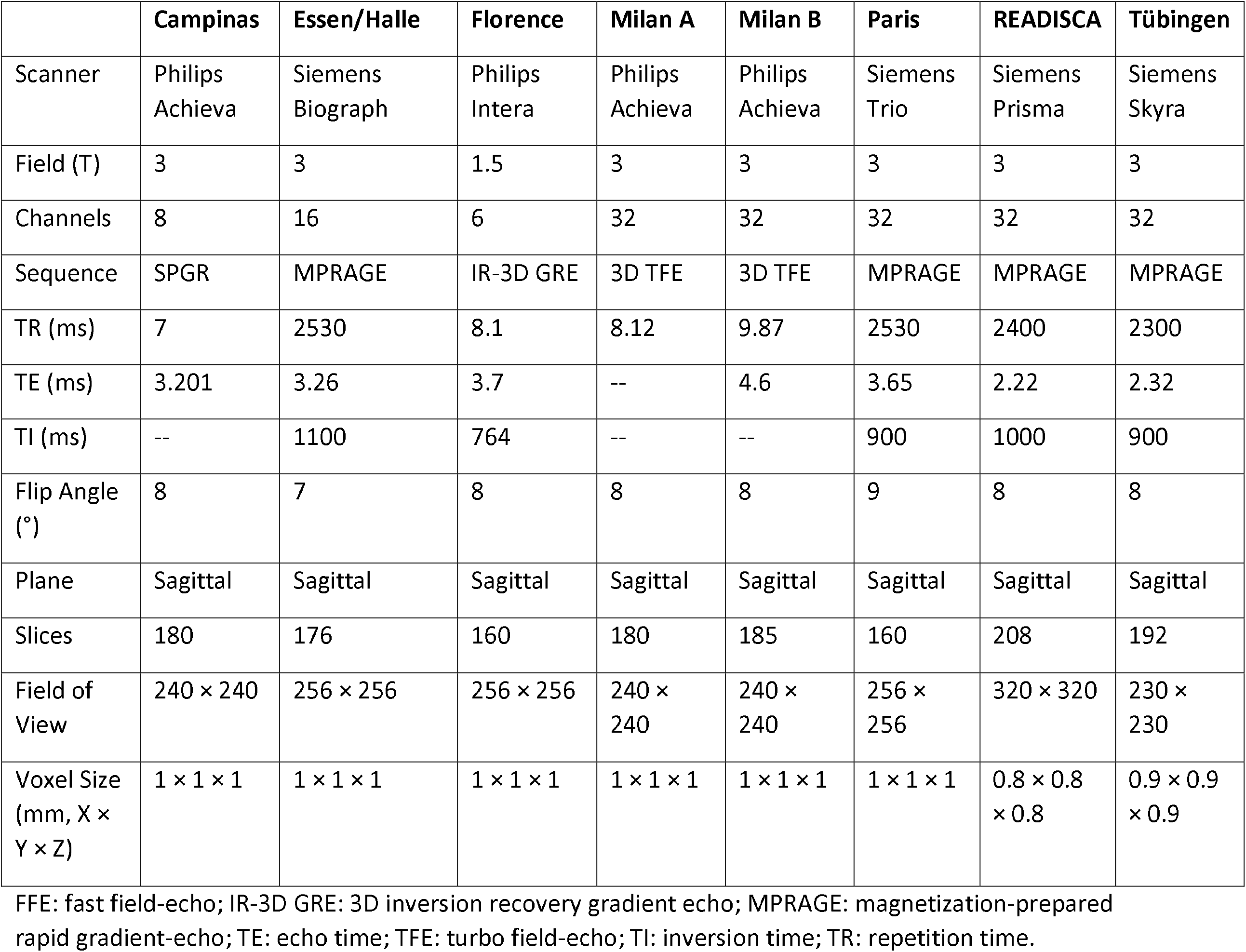
Summary of imaging protocols as reported by each site.

**Supplementary Table S2.** Demographic statistics for the four cohorts of Duration and CAG repeat length portrayed in Figure 6. Pairwise comparisons performed with the Kruskal-Wallace non-parametric test with Bonferroni correction for multiple comparisons.

|  | Duration ≤ 7 years (L) | Duration > 7 years (H) |  |
| --- | --- | --- | --- |
| **CAG Repeats ≤ 44 (L)** | 8 (5-9.5) | 14.5 (9.25-18.5)* | SARA Score |
|  | 3 (2-6) | 15 (9.5-19.5)* | Duration |
|  | 41 (39-43) | 42 (41-44) | CAG Repeats |
| **CAG Repeats > 44 (H)** | 11.5 (8.5-17)* | 13.5 (11-21.5)* | SARA Score |
|  | 5 (4-6)† | 12 (11-14)*‡ | Duration |
|  | 49 (46-52)*† | 47 (46-51)*† | CAG Repeats |

All values are expressed as median (interquartile range).
\* significantly different from Duration L/CAG L
† significantly different from Duration H/CAG L
‡ significantly different from Duration L/CAG H

**Supplementary Table S3.** Linear model statistics from cerebrocerebellar covariance analysis.

|  | Model Fit |  |  | *cerebellum* |  | *Group* |  | *ICV* |  | *cerebellum × Group* |  |
| --- | --- | --- | --- | --- | --- | --- | --- | --- | --- | --- | --- |
| | $F_{4,240}$ | $p$ | Adj. $R^2$ | $t$ | $p$ | $t$ | $p$ | $t$ | $p$ | $t$ | $p$ |
| GM only | **59.91** | **< 0.001** | **0.491** | **4.084** | **< 0.001** | 1.383 | 0.168 | **9.940** | **< 0.001** | -1.492 | 0.137 |
| WM only | **72.80** | **< 0.001** | **0.541** | **6.434** | **< 0.001** | **3.205** | **0.002** | **7.124** | **< 0.001** | **-2.933** | **0.004** |
| GM + WM | **93.09** | **< 0.001** | **0.602** | **6.159** | **< 0.001** | **2.819** | **0.005** | **10.07** | **< 0.001** | **-2.710** | **0.007** |

Adj. $R^2$ : Adjusted $R^2$ ; ICV: intracranial volume; GM: grey matter; WM: white matter

